## Supplement Table for "Low seroprevalence of Ebola virus in health care providers in an endemic region (Tshuapa province) of the Democratic Republic of the Congo"

**Supplement:** Seroprevalence with cut off obtained from literature for different (combinations of) antibodies against Ebola virus antigens as measured by the Luminex or FANG ELISA in Health care providers from Boende, DRC.

| | Antigen | | Positives | Seroprevalence | Age /year | M *vs* F | Direct Contact with patients: Direct vs indirect | Working Hospital vs elsewhere | Experienced Ebola outbreak/patients vs others |
|---|---|---|---|---|---|---|---|---|---|
| | | **Cutoff** | **n (N)** | **% (95% conf. Int.)** | **p-value** | **p-value** | **(p-value)** | **(p-value)** | **p-value** |
| **FANG ELISA** | **GP-EBOV-m** | 607 EU/ml | 49 (694) | 7 | 0.73 | 0.35 | 0.24 | 0.67 | 0.61 |
| | **GP-EBOV-m** | 381 MFI/100 beads | 104 (698) | 14 | **0.005** | 0.22 | 0.05 | 0.81 | 0.24 |
| | **GP-EBOV-k** | 501 MFI/100 beads | 89 (698) | 13 | **0.02** | 0.81 | 0.05 | 0.99 | 0.45 |
| | **VP40-EBOV-m** | 580 MFI/100 beads | 69 (698) | 10 | 0.14 | 0.11 | 0.41 | 0.35 | 0.16 |
| **Luminex** | **NP-EBOV-m** | 950 MFI/100 beads | 8 (698) | 1 | 0.54 | 0.57 | 0.31 | 0.16 | 0.08 |
| | **GP-EBOV-m+NP-EBOV-m** | C1 | 0 (698) | 0 | | | | | |
| | **GP-EBOV-m+VP40-EBOV-m** | C2 | 19 (698) | 3 | 0.92 | 0.37 | 0.91 | 0.38 | 0.13 |
| | **NP-EBOV-m+VP40-EBOV-m** | C3 | 2 (698) | 0.2 | | | | | |
| **Luminex and FANG ELISA** | **GP-EBOV-m** | C4 | 6 (694) | 0.8 | | | | | |

*cutoff (C) represent values obtained from literature and previous studies

C1= 381 MFI/100 beads for GP-EBOV-m and 580MFI/100 beads for NP-EBOV-m

C2=381 MFI/100 beads for GP-EBOV-m and 950 MFI/100 beads for VP40-EBOV-m

C3= 950 MFI/100 beads for NP-EBOV-m and 580 MFI/100 beads for VP40-EBOV-m

C4= 607 EU/ml and 381MFI/100beads for GP-EBOV-k and GP-EBOV-m
